# Adapting a Regulation of Craving Magnetic Resonance Imaging Task to Generate Functional Repetitive Transcranial Magnetic Stimulation Targets for the Ventromedial and Dorsolateral Prefrontal Cortex in Treatment-Seeking Participants with Cannabis Use Disorder

**DOI:** 10.64898/2026.06.04.26353616

**Authors:** Andrew Geoly, Daniel M. McCalley, Wiebke Struckmann, Azeezat Azeez, Brendan Wong, Bohye Kim, Seigo Ninomiya, Sumaiya Ahmed, Jane P. Kim, Aimee L. McRae-Clark, Brett Froeliger, Gregory L. Sahlem

## Abstract

**Background:** Repetitive Transcranial Magnetic Stimulation (rTMS) is a promising treatment across addictive disorders including Cannabis Use Disorder (CUD). Targeting incentive-salience circuitry via the ventromedial prefrontal cortex (vmPFC) and central-executive circuitry via the left dorsolateral prefrontal cortex (LDLPFC) are both promising treatment approaches; however, to date structural targets have predominated whereas functional targeting may allow for more precision. In this pilot trial we adapted a functional Magnetic Resonance Imaging (fMRI) Regulation of Craving (ROC) task to generate fMRI-based rTMS targets in the vmPFC and LDLPFC.

**Methods:** We recruited treatment-seeking participants with moderate or severe CUD as a part of an open-label trial and administered an adapted ROC-task during fMRI following 24-hours of cannabis abstinence. We identified sub-portions of maximal activation of the LDLPFC when participants thought of long-term consequences of cannabis use (Later) and of the vmPFC when participants thought of short-term positive aspects of cannabis use (Now). We hypothesized that our task would generate acceptable rTMS targets in >66% of baseline fMRI scans.

**Results:** A total of 20-participants enrolled in the trial (50%F, age=33.3±9.8) and completed the baseline fMRI. The adapted ROC-task elicited group level activation in the LDLPFC and precuneus in the Later>Now and in the bilateral vmPFC, ACC, and striatum in the Now>Later contrast. Acceptable functional targets resolved in both the vmPFC and LDLPFC in 19 of 20 participants (one participant did not tolerate MRI).

**Conclusions:** The adapted ROC-task elicits activation in incentive salience and central executive circuitry and can feasibly generate rTMS targets when using a cluster selection algorithm.

## Introduction

Cannabis Use Disorder (CUD) is an increasingly problematic substance use disorder, with a high prevalence and substantial morbidity in the United States. To date, there is no approved or standard of care pharmacotherapy for CUD, and behavioral therapy is not effective for all patients^1^. Subsequently, there is a need to develop and test novel treatments for CUD. One such novel treatment option is repetitive Transcranial Magnetic Stimulation (rTMS), which has shown promise as a treatment for CUD^2,3^ and a variety of substance use disorders^4^. To date, two predominant rTMS strategies have emerged: 1) delivering rTMS to the left dorsolateral prefrontal cortex (LDLPFC) to enhance executive/inhibitory control or 2) delivering rTMS to the ventromedial prefrontal cortex (vmPFC) to reduce limbic drive toward substance use^5,6^. While both of these strategies have shown promise in reducing substance use, there is a gap in our knowledge regarding how an individual patient should be matched to an rTMS target. One strategy to select an rTMS target may be to leverage individualized functional Magnetic Resonance Imaging (fMRI) data. For example, preliminary data suggests that, among patients with substance use disorders, delivering rTMS to a brain region with high functional activity enhances its efficacy^7^.

While the development process for fMRI-guided rTMS for depression is a useful blueprint upon which to build a similar strategy in addiction, there are several refinements in experimental approach which may be more germane to substance use. For example, in rTMS for MDD, functional connectivity to the subgenual cingulate cortex has been used to derive precise, cortical rTMS targets^8,9^. In substance use disorders, however, it is not clear that a single, subcortical brain region and its associated cortical circuitry drive symptoms. Given that rTMS is a relatively focal treatment, and different cortical targets have different sub-cortical downstream effects^10^, using connectivity-based MRI targeting may not always precisely target the correct down-stream subcortical region for a given individual.

Further, in MDD, rTMS targets are derived from fMRI data obtained during rest (e.g. a resting-state scan). In substance use disorders, however, task-based cognitive^11^ and drug cue-reactivity tasks^12^ are symptomatically germane, linked to relapse, and can reliably probe brain dysfunction on an individual basis. The Regulation of Craving (ROC) task^13^, a well-established fMRI paradigm in addiction research, can elicit distinct activation in central-executive and limbic circuitry when probing processes related to DLPFC-mediated executive function and vmPFC-mediated drug cue-reactivity, respectively. Being able to elicit activation in these two important cortical targets, and their corresponding down-stream target regions using a single fMRI-task makes the ROC task a promising method of generating rTMS treatment targets and providing brain-state to personalize treatment.

In order to test the feasibility and preliminary benefit of using an adapted ROC-task for rTMS targeting, we conducted a pilot open-label randomized-controlled trial that built upon our previous work. We developed an MRI paradigm to use the ROC-task to generate rTMS treatment targets for the vmPFC and LDLPFC and to preliminarily determine if we could elicit expected network activation to quantify the magnitude of activation and potentially develop a biotyping paradigm in the future. We hypothesized we would be able to feasibly generate rTMS targets in both target brain regions, and a-priori set a threshold of 66% of participants as the bar for feasibility (reasoning that this was the minimum number of functional targets needed to justify the added cost and labor of using MRI). We further tested clinical and fMRI outcomes of delivering a course of open-label rTMS to each of the treatment targets in a randomized fashion, and these results will be reported separately.

## Methods

### Overview, Recruitment, Screening, and Behavioral Assessments

We recruited outpatient, treatment-seeking participants with CUD through media advertisements and through addiction medicine clinical referrals in the area surrounding Palo Alto, California. All participants were recruited through a parent pilot randomized controlled trial, NCT05720312, testing the preliminary efficacy of either high-frequency rTMS applied to the LDLPFC, or low-frequency rTMS applied to the vmPFC. The parent trial was registered with clinicaltrials.gov, approved by the Stanford University Institutional Review Board, and was conducted in accordance with the principles outlined in the Declaration of Helsinki. All participants signed informed consent prior to beginning study procedures.

Participants underwent a standard screening and enrollment visit which included a clinical history and exam, assessment using the Quick Structured Clinical Interview for the 5^th^ edition of the Diagnostic and Statistical Manual of Mental Disorders^14^ which included the criteria for Cannabis Use Disorder. Participants further were evaluated using the Time-Line Follow-Back^15^, the 3-item Craving Scale^16^, the Marijuana Problem Scale^17^, and the Cannabis Withdrawal Scale^18^. Participants were enrolled in the study if they were older than 18 years old, they met criteria for moderate or severe cannabis use disorder, expressed interest in cannabis cessation or reduction, and had sufficient intelligence and command of the English language to complete informed consent procedures and study related assessments. Participants were excluded if they were pregnant or breastfeeding; met criteria for any other moderate or severe substance use disorder (other than tobacco use disorder); had changes in psychoactive medications in the 4-weeks preceding enrollment; had a history of bipolar affective disorder, schizophrenia, dementia, active suicidal ideation, or a suicide attempt within the past 6-months; had any contraindications to rTMS^19^ or MRI^20^; or had other unstable medical, neurologic, or psychiatric disorders.

Participants who enrolled in the trial underwent a baseline MRI visit following 24-hours of abstinence from cannabis and other substances verified by both self-report and saliva drug testing (12-panel now, Boynton Beach, Fl). Prior to the MRI visit they underwent standard assessments including cannabis withdrawal^18^ and craving^16^ over the past 24-hours. Following these assessments, participants were trained on the adapted ROC-task as described below and similar to the training paradigm developed by Kober and colleagues^13^.

### Task Development and MRI scanning paradigm

#### Task paradigm

This study used an adapted version of the ROC task described by Kober and colleagues^13^ for rTMS targeting for CUD. During the task, participants viewed cannabis cues personalized based on the participants most frequent route of cannabis administration (joint, bong, vape/concentrate), or matched neutral images of toothbrushes^21^. Prior to viewing each cannabis cue, participants were instructed to either think about their personal short-term benefits of using cannabis via a ‘Now’ prompt, or the potential long-term negative consequences of continued cannabis use at their current rate using a ‘Later’ prompt. Prior to viewing neutral images participants were instructed to not think of anything in particular using a “Relax” prompt. As shown in Figure-1, each trial began with a fixation cross (jittered around 4-seconds), followed by an instruction prompt as described above (‘Now’, ‘Later’, or ‘Relax’) for 2-seconds. This was followed by the cannabis or neutral image for 6-seconds, followed by another fixation cross (jittered around 3-seconds). Finally, participants rated their current craving, with the craving indicated on a scale from 1-to-5 (with 1=none, 2=mild, 3=moderate, 4=severe, and 5=extreme craving) for 3-seconds. Each task run contained a total of 40-trials, consisting of 20-cannabis images (10 ‘Now’ and 10 ‘Later’) and 20 neutral images, resulting in a block length of approximately 12-minutes. A neutral condition was presented every other trial to minimize unexpected shifts between stimulus types, ensuring consistent task engagement, while the order of the ‘Now’ and ‘Later’ conditions was pseudorandomized. The task consisted of up to three blocks, thus up to 36-minutes total, each separated by a short break. In the first and third task block, drug image stimuli were based on the preferred cannabis usage modality by the participant, as established during the screening process, i.e. joint, bong, or vaping, to better target the individual craving response. In the second task block, drug image stimuli were a combination of all three usage modalities. We elected to administer up to three blocks of the task (up to 36-minutes) due to the exploratory nature of this trial and to ensure that there was sufficient brain signal evoked by the task stimuli to generate rTMS-targets.

**Figure-1.**
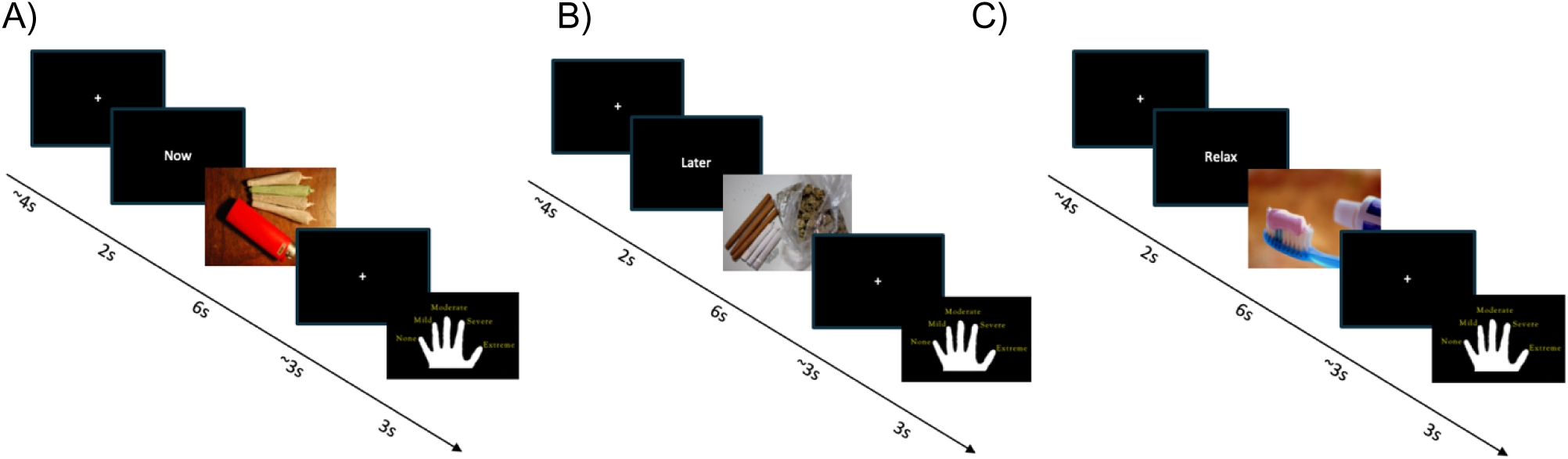
Basic Task structure: general organization of the Regulation of Craving task (Figures 1A-C), which includes trials of ‘NOW’ in Figure 1A, ‘LATER’ in Figure 1B, and Neutral in Figure 1C.

#### Task training

An initial task training session was conducted outside of the scanner, with a different set of images for training and scanning. Participants were shown 13 trials in the training session (4-Later, 3-Now, 6-Neutral) which could be repeated as needed. Similar to Kober and colleagues, once participants reported the ability to consistently think about the short-term and long-term consequences of their cannabis use, and follow all task instructions, the scanning session was started. In this pilot trial each participant needed only a single training session to report competence in completing the task.

#### MRI scanning parameters

Participants were scanned in a GE Discovery MR750 scanner (GE Medical systems, Chicago, Illinois, USA) with a Nova 32-channel head coil. Functional images were acquired with a T2*-weighted EPI BOLD sequence with a TR of 1,600ms, TE of 30ms, flip angle of 60°, 116 x 116 in-plane matrix, field of view of 23.2cm, and 72 2mm-slices in an interleaved order. Anatomical images were acquired with a T1-weighted sequence with a TR of 6.388ms, TE of 2.624ms, flip angle of 12°, 256 x 256 in-plane matrix, field of view of 23.0cm, and 0.9mm-slices covering the entire brain of the participant. Phase Reverse scans were acquired for distortion correction.

### MRI pre-processing, Modeling, and Target Generation

#### Pre-Processing

A detailed description of the preprocessing steps, which were performed using fMRIPrep^22^, can be found in the Supplement. In short, anatomical preprocessing included bias field correction, skull stripping, tissue segmentation, surface reconstruction, and nonlinear normalization to MNI standard space. Functional preprocessing involved motion correction, susceptibility distortion correction, co-registration to anatomical images, and spatial smoothing (6mm FWHM). Noise reduction was performed using ICA-AROMA for motion artefacts and CompCor for physiological noise, with confound regressors including framewise displacement (>0.5mm), DVRAS, and global signals. Preprocessed BOLD data were resampled into standard space using Lanczos interpolation.

#### General Linear Models

First-level general linear models (GLM) were constructed using the Statistical Parametric Mapping (SPM12) toolbox implemented in MATLAB (R2018a). Native-space, de-skulled, and grey-matter masked BOLD EPI data were input into each model. The model was fit with 11 task regressors (‘Now’, 3 regressors: prompt, image cue, and relax; ‘Later’, 3 regressors: prompt, image cue, and relax; Hand-pad ratings, 3 regressors: following now cue, following later cue, and following neutral cues; Fixation crosses, 2 regressors), 6 motion regressors (X, Y, Z, pitch, roll and yaw), a constant regressor, and a high pass filter cutoff of 100 seconds. Motion-adjusted contrast maps were created for each task condition. Primary contrasts of interest for whole-group, second level analyses were defined as ‘Now>Later’ (response to Now Cue > Later Cue) and ‘Later>Now’ (response to Later Cue > Now Cue). For the second level, whole-group analysis, we used a voxel-wise threshold, p<0.05, cluster-level threshold, p<0.05, Family-wise error corrected).

#### Target generation

Custom-made targeting masks were constructed in MNI space to generate targets for the L-DLPFC and the bilateral vmPFC (See Figures 2-3). The L-DLPFC search space was chosen based on our group’s previous work delivering rTMS to this area specifically^23^. We chose a bilateral vmPFC mask given the high degree of activation variability in previous cue-reactivity trials showing that there may not be a clear lateralization (maximally activated clusters may be present in the left, right, or along the midline of the vmPFC)^7,24^. As such, a bilateral vmPFC search space accounted for the high degree of inter-individual variability and optimized our personalized targeting approach. Standard MNI search spaces were normalized to the participant’s native T1w anatomical space using a custom in-house co-registration script that performed N4 bias field correction on the T1w image with ANTs, skull-stripping with *mri_synthstrip* from Freesurfer (7.4.1), affine and nonlinear b-spline diffeomorphic registration and mask back-transformation into native T1w with ANTs. Finally, T1w search spaces were resampled into the dimensionality of 1^st^-level SPM Z-contrast maps for further clustering analyses. Next, whole-brain Z-statistic maps from each task run were masked to the L-DLPFC and vmPFC search spaces in parallel. The resulting maps were then smoothed using a Gaussian filter with a FWHM of 2mm, as implemented in FSL. For each participant, all three task runs were evaluated independently and also averaged to create a combined Z-map.

**Figure 2.**
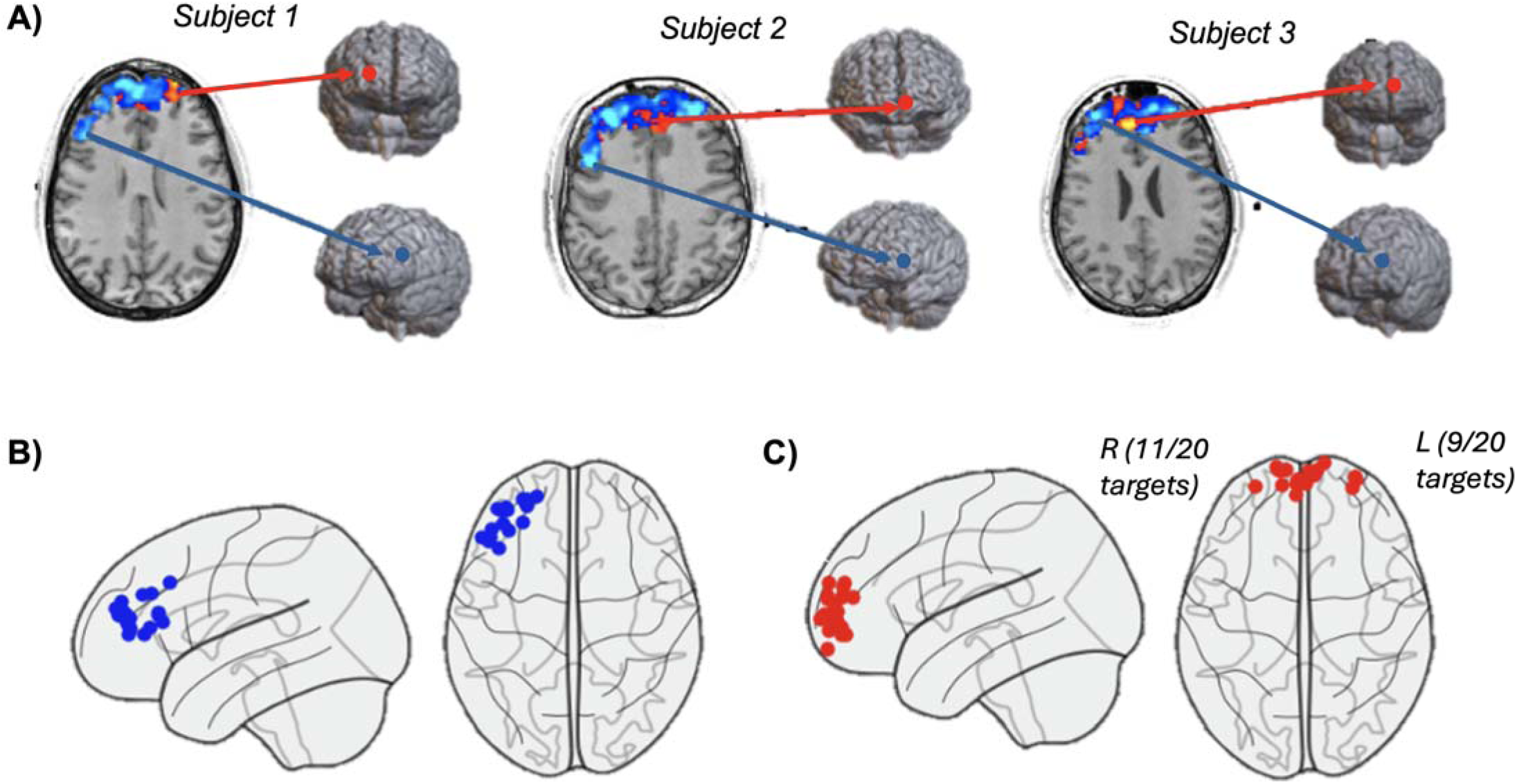
rTMS target generation in 3 example subjects (A) and topography of group-level rTMS targets (B) and (C) Figure 2: (A) Statistical parametric map of ‘Now’ (Red) > ‘Later’ (Blue) cues across L-DLPFC and bilateral vmPFC targeting search mask ROIs (Z = –1:1) and corresponding peak target locations for three example participants. Target positions mapped to MNI coordinate space in sagittal (left) and axial (right) views for L-DLPFC search mask (B) and bilateral vmPFC (C). 11/20 vmPFC personalized targets generated to the right hemisphere and 9/20 to the left hemisphere. Interindividual variability in positional spread is evident across both search masks with greater variability present in the bilateral vmPFC.

**Figure 3:**
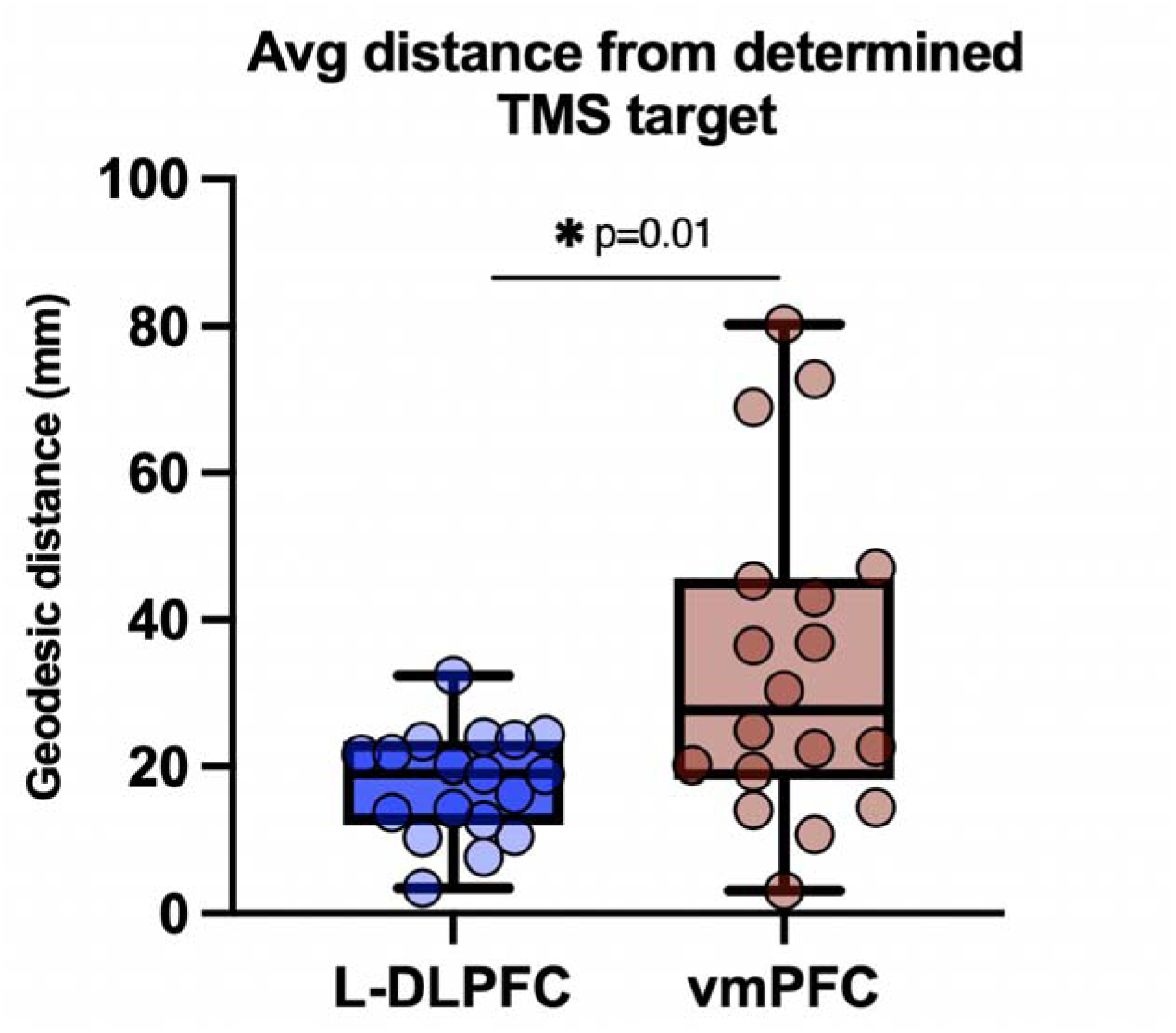
Spatial reliability of fMRI guided TMS targets within the L-DLPFC and vmPFC. Figure 3: Within-subject average geodesic distance between the selected TMS target across 3 task runs and the remaining generated cortical targets. Spatial spread of targets was significantly lesser within the L-DLPFC than the vmPFC (t=2.8, p=0.01). Each circle represents within-subject average geodesic distance from the selected target to the targets generated from each other run of the task. Mid lines of the box plots represent mean.

**Figure 4:**
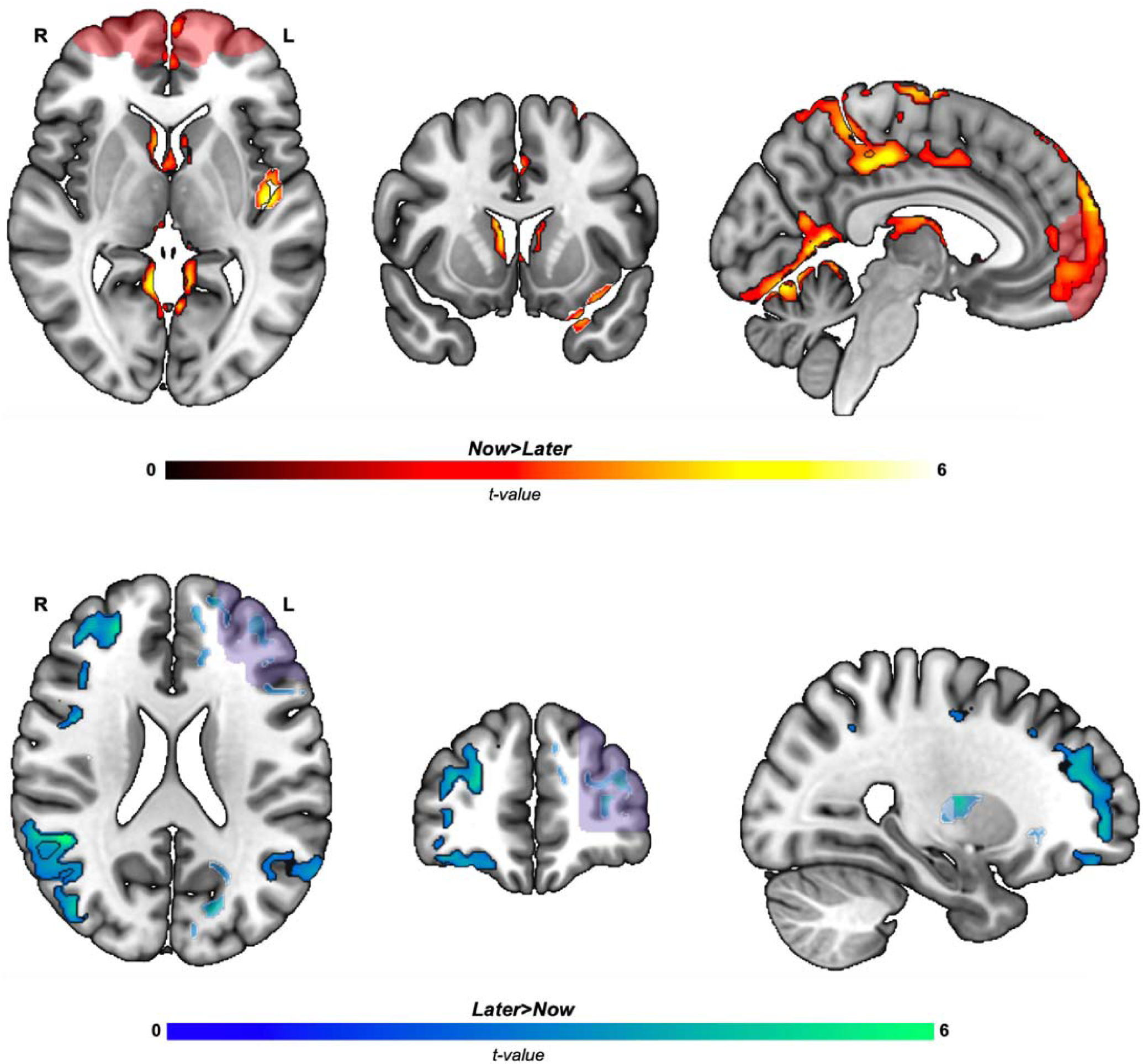
Elevated fMRI BOLD signal in response to ‘NOW’ and ‘LATER’ cues, relative to one another. Figure 4: Top Panel; Elevated fMRI BOLD signal in response to ‘NOW’ relative to ‘LATER’ cues (r d). The search mask used for TMS targeting is visible in a faded red color. Bottom Panel; Elevated fMRI BOLD signal in response to ‘LATER’ relative to ‘NOW’ cues (blue). The search mask used for TMS targeting is visible in a faded blue color. Regions outlined in black survive multiple comparison correction (voxel-wise threshold, p<0.01, Family-wise error multiple comparison corrected), regions outlined in white do not survive strict, cluster-level multiple comparison correction but were significant at the voxel-level (voxel-wise threshold, p<0.01, cluster threshold, p<0.05, uncorrected). Color-bar represents magnitude of t-value difference etween contrasts. (L=left, R=right).

To identify statistically relevant clusters within each masked and smoothed map, we first established a subject-specific cluster inclusion threshold based on the empirical distribution of Z-values. Voxels exceeding the 95th percentile of this distribution were retained for cluster formation for each respective contrast map (e.g., ‘Later > Now’ for the L-DLPFC and ‘Now > Later’ for the vmPFC). Clustering was performed using FSL’s cluster tool, configured to detect clusters with a minimum size of 15 contiguous voxels, 6-connected voxel adjacency, and voxel-wise Z-values exceeding the 95^th^ percentile.

Clusters generated by FSL were then evaluated using custom MATLAB scripts to quantify cluster size (number of voxels), mean Z-statistic, coefficient of variation, and cluster kurtosis. A composite score was calculated for each cluster using a weighted sum of these metrics favoring larger clusters with higher-magnitude Z-values and penalizing clusters with high coefficient of variation and kurtosis. The cluster with the highest composite score was selected as the primary target for that map, and the corresponding region was saved as a binary NIfTI image. Finally, given a total of four clusters were created per subject (corresponding to the three tasks runs and their overall average) a review process was conducted by the research team to assess the selected cluster’s spatial location, shape, and consistency across runs. Individual cluster metrics and overlays were inspected in each run (run 1, run 2, run 3, and total average). If necessary, additional manual thresholding or refinement was applied to ensure anatomical and statistical validity of the selected target.

To quantify the spatial stability of our targets across runs, the center of gravity of each cortical target was warped into MNI152 space and the geodesic distance was computed along the cortical surface mesh. Each target coordinate set from each subject was projected to the nearest surface vertex using a nearest-neighbor approach. Within each subject, the vertex location for the determined TMS treatment target served as the reference point. Dijkstra’s shortest-path algorithm was applied to calculate the geodesic distance in millimeters from the reference vertex to the remaining generated TMS targets. Each measure of geodesic distance was then averaged within-subject and within search space (e.g. L-DLPFC or vmPFC). The relative difference in distance across targets was compared within the L-DLPFC and vmPFC with a paired-samples t-test.

### Relationships between fMRI and Behavioral Assessments

In the original use of this task for tobacco use disordered participants, Kober and colleagues related the degree of hand-pad craving to the condition (Now or Later), and the degree of activation in the ventral striatum (VS) and the left dorsolateral prefrontal cortex (LDLPFC). They found that hand-pad craving was higher in the Now runs than the Later runs, and the percent less craving the participant reported in the Later relative to Now runs positively correlated with L-DLPFC activation and negatively correlated with VS activation. Specifically, they generated percent signal change (both positive and negative) in the contrast Later > Now, performed a correlation analysis with the percent change between Later and Now ratings of craving (%craving was lower in Later runs than Now runs), and determined the correlation relationship between DLPFC and VS with percent signal change. They then performed a pathway analysis to see if VS deactivation mediated the relationship between DLPFC activation and percent-craving reduction using the hand-pad.

We attempted a replication of this finding using their methodology to the best of our ability. We first used Marsbar to extract the average BOLD signal during the contrast of Later > Now in the LDLPFC and bilateral ventral striatum (Oxford-GSK_Imanove connectivity atlas) in the combined activation map. We then calculated the average hand-pad rating following each type of rating (Now, Later, Neutral). We then calculated the percent decrease in hand-pad craving rating between the Later and Now trials and used Spearman’s rank correlation test to determine if the extracted BOLD signal in the VS and LDLPFC correlated with the percent decrease in craving between Later and Now trials. Finally, we aimed to perform the pathway analysis using the standard three-variable path model, similar to the Kober et al analysis description. Given the limited sample size and focus on generating rTMS targets in this experiment, we limited our behavioral analysis to this attempted replication of Kober’s findings.

## Results

### Participant Characteristics (Table-1)

We assessed a total of 30 potential participants for entry into the trial, of which twenty-two were initially found eligible and enrolled. One of the initially enrolled participants never attended their scheduled MRI-visit, and a second was later found to meet an ineligibility criteria upon medical records review. There subsequently was a final sample of twenty enrolled participants. One participant could not tolerate the MRI environment due to claustrophobia. Subsequently, nineteen of the twenty enrolled participants completed the Targeting MRI visit and contributed to the MRI data.

### rTMS Target Generation (Figure-2), Spatial Reliability (Figure-3) and Baseline fMRI Characteristics (Figure-3, Supplemental Table-1, Supplemental-Figure-1, and Supplemental-Table-2)

We were able to generate individualized TMS targets within both the vmPFC and LDLPFC in 19 of the participants who were able to tolerate MRI (95% of the original sample, Figure 1).

On the group level, there were three primary clusters wherein fMRI BOLD signal was elevated in response to ‘Now’ relative to ‘Later’ cues, (k_1_=3,996, k_2_=826, k_3_=2,101) with local maxima in the pre and post central gyri (bilateral), the anterior and middle cingulate gyri (left), the precuneus (bilateral), and the ventral and dorsomedial prefrontal cortices (left and right), the lingual gyrus (bilateral), and the caudate (bilateral) (voxel-wise p<0.01, Family-wise error multiple comparisons corrected). At a more liberal statistical threshold (voxel-wise p<0.01, cluster-wise threshold, p<0.05, uncorrected), there was an additional local maximum in the left posterior insula.

There were 3 primary clusters wherein fMRI BOLD signal was elevated in response to ‘Later’ relative to ‘Now’ cues (k1=2,572, k2=1,875, k3=1,535), with local maxima in the angular gyrus (bilateral), dorsolateral prefrontal cortex (right), Broca’s area (right), and the ventrolateral prefrontal cortex (right) (voxel-wise p<0.01, Family-wise error multiple comparisons corrected). At a more liberal statistical threshold (voxel-wise p<0.01, cluster-wise threshold, p<0.05, uncorrected), there was an additional local maximum in the left dorsolateral prefrontal cortex.

### Baseline Behavioral Characteristics and Relationship with fMRI Characteristics

MRI hand-pad craving was highest in the Now runs (3.3 ± 0.9), followed by the Later runs (2.9 ± 0.9), followed by the Neutral runs (2.3 ± 0.8), with the average craving 8% lower in the Later runs than Now runs (Wilcoxon signed rank test; p = 0.0005). There was not a significant relationship between the percent craving reduction in Later runs relative to Now runs and LDLPFC (rho = –0.03; *p*=ns) and VS (rho = –0.26; *p*=ns). There was no association between the LDLPFC and the VS, which is an eligibility criterion for the mediation analysis; therefore, we did not proceed with the mediation analysis.

## Discussion

There is substantial momentum to develop personalized, fMRI-guided rTMS approaches to enhance clinical outcomes beyond traditional rTMS methods. While there have been several interesting findings which demonstrate delivering rTMS to a functionally engaged target may boost treatment outcome, these data have been retrospective in nature. Thus, there is a need to develop a rigorous pipeline to generate fMRI-guided rTMS targets in populations with substance use disorder. Here, in this small proof of concept trial, we demonstrate that functional MRI targets can be feasibility generated in 19/20 cases within the vmPFC and DLPFC and we replicate the findings of Kober and colleagues, finding that in Later > Now runs, central executive circuitry activated including the LDLPFC, and in Now > Later runs, incentive salience circuitry activated including the vmPFC, ACC, and striatum^13^. We did not replicate the relationship between task activation and behavioral variables, though we did replicate the finding that craving was reduced during Later runs relative to Now runs.

To date, to our knowledge, all published treatment trials using rTMS to treat addictive disorders have used either scalp-based or structural-MRI-based targeting. Though a well-established methodology, structural-anatomical imaging misses functional-anatomical information that may have important ramifications for treatment effects^7,11,25^. Specifically in recent cue-reactivity work there was substantial variability in activation patterns in both participants with cocaine and alcohol use disorders^24^, and in a recent clinical trial using rTMS to treat alcohol use disorder overlapping rTMS-induced electrical fields and activation pattern explained much of the treatment effect^7^. In our trial we were able to generate robust activation maps and use our cluster selection algorithm to isolate rTMS targets in all of the participants who were able to tolerate MRI. The ability of this task and targeting algorithm to generate targets in 95% of the study group (and 100% of those undergoing MRI) demonstrate the feasibility of this targeting method (far exceeding the 66% a priori cutoff we selected), and may make this a clinically translatable paradigm should there be beneficial behavioral effects following the application of a course of treatment.

Across subjects, there was a relatively high degree of spatial variability in the target location within the LDLPFC and vmPFC targeting masks, as can be seen in Figure 2. That said, within subjects, there was a high degree of consistency between each target calculated across three task runs and their weighted combination. For example, the average distance between the selected TMS target and the not-selected targets was 1.7cm and 3.4cm, within the LDLPFC and vmPFC, respectively. Given others have published that most TMS figure-of-eight coils produce an electrical field with a breadth of about 5cm, this indicates that in nearly all of our subjects, delivering TMS to the chosen target would reach the remaining targets which were not selected. There is notably a wider spread of targets both within and across subjects within the vmPFC, however, we note that the search space for that specific mask extended to both brain hemispheres, as there is no precedent clear^7,247,26^. Taken together, these data suggest the techniques used within this manuscript can generate reliable TMS targets within a given participant, however future work may explore whether the TMS target determined for a given individual is stable across serial MRI scans.

We replicated two key findings of Kober and colleagues in the development of the ROC task for substance use disorders. First, we find robust activation of regulatory structures (specifically the DLPFC) during the Later relative to Now, and Neutral trials and in incentive-salience structures in the Now relative to Later, and Neutral trials (Figure-3 and Supplemental-Figure-1). Second, we also find that hand pad rating was lower in Later trials relative to Now trials. We did not, however, replicate the relationship between neural activation and hand-pad ratings. Reasons for this disparate finding are unclear, however of note this is a different population (with well demonstrated cognitive dysfunction)^26,27^. Further the study sample was in some degree of cannabis withdrawal (as evidenced by an increase in the cannabis withdrawal scale score relative to screening scores with the required 24-hours of cannabis-abstinence prior to MRI) which may have resulted in a ceiling effect of craving. Both of these variables differ from the tobacco use disordered population in the original cohort. Of note, the present cohort also had a small sample size which limited power to detect such a difference, though it was of a similar size to the sample previously reported by Kober and colleagues.

## Limitations

Though this investigation had a number of strengths including rigorous screening and evaluation methods to include a treatment seeking cohort with CUD, a strong biologically informed task-paradigm development, and state of the art fMRI techniques; there are several limitations that warrant mention. First, even though we were able to generate targets for all individuals completing MRI, our study sample was relatively small, and so we may not be able to generate targets for all participants in a larger sample. Secondly, though we chose our targeting masks based on previously published literature, targeting was completed within only discreet masks as opposed to generating mask-agnostic targets. Finally, all scans occurred on a single day and there was no ability to determine test-retest reliability. Despite these limitations this trial represents a first step in the development of more precision paradigms with the potential to improve the strength and reliability of clinical effects of rTMS in cannabis use and addictive disorders more broadly. Future work should address the present limitations, including a larger sample, as well as test-retest evaluations, in the context of a clinical trial.

**Table-1.** Baseline and demographic characteristics of the study sample: All values are reported ± Standard Deviations. Cannabis use variables are reported for the 28-days prior to the screening and enrollment visit.

| Sample Demographics: N=20 participants |  |
| --- | --- |
| Age (Years) | 33.3 ± 9.8 |
| Biological Sex | 50% Male, 50% Female |
| Ethnicity (NIH Criteria) | 70% non-Hispanic<br>30% Hispanic<br>0% Unknown |
| Race (NIH Criteria) | 15% Caucasian<br>20% Black / African American<br>15% Other<br>5% Asian<br>5%Native Hawaiian/Other pacific Islander<br>40% More than one<br>0% Native American |
| % ≥4-year college degree | 50% |
| Marital Status (%married) | 10% |
| Employment (%full time) | 50% |
| % Daily tobacco users | 20% |
| Cannabis Use Characteristics: |  |
| Age of first cannabis use | 15.4 ± 2.4 |
| Age of onset of >weekly cannabis use | 18.9 ± 4.8 |
| Number of prior quit attempts | 4.1 ± 5.9 |
| Longest previous period of abstinence (months) | 8.9 ± 17.1 |
| Number of DSM-5 CUD categories met | 9.7 ± 1.0 |
| Most common cannabis use method (%): | 35% Using a 'bowl, bong, pipe, or other similar'<br>35% Using a 'blunt / Joint / spliff'<br>30% Using a vaporizer, dabs, waxes, or concentrates<br>0% Two or more methods approximately equally |
| Marijuana Problem Scale Total Score | 14.7 ± 6.3 |
| Study goal: | 0% Minimal Reduction in use<br>5% Partial Reduction (use less per use-session or fewer times per day, but still 5+ days per week)<br>5% Moderate Reduction (cannabis <5 days / week)<br>25% Substantial Reduction (cannabis <3 days / week)<br>25% Near abstinence (use cannabis < once per month)<br>40% Abstinence (true-quit attempt) |
| Cannabis Withdrawal Scale symptom score with ad libitum use | 58.7 ± 30.5 |
| Cannabis Withdrawal Scale symptom score at 24-hours of abstinence at the MRI visit | 82.6 ± 26.3 |
| Days of cannabis use (previous 28-days) | 27.7 ± 0.8 |
| Average number of cannabis use sessions per day (previous 28-days) | 6.1 ± 3.6 |

| MRI Hand-pad Craving Values |  |
| --- | --- |
| Craving Later runs | 2.9 ± 0.9 |
| Craving Now runs | 3.3 ± 0.9 |
| Craving Neutral runs | 2.3 ± 0.8 |
| 3-Item Craving Score at Screening (ad libitum cannabis use) | 21.7 ± 4.5 |
| 3-Item Craving Score at Screening (24-hours of abstinence) | 23.2 ± 3.7 |

DSM-5 CUD: Cannabis Use Disorder Criteria from the Diagnostic and Statistical Manual for Mental Disorders

## Supplemental Methods, Figures and Tables

**Supplemental-Figure-1.**
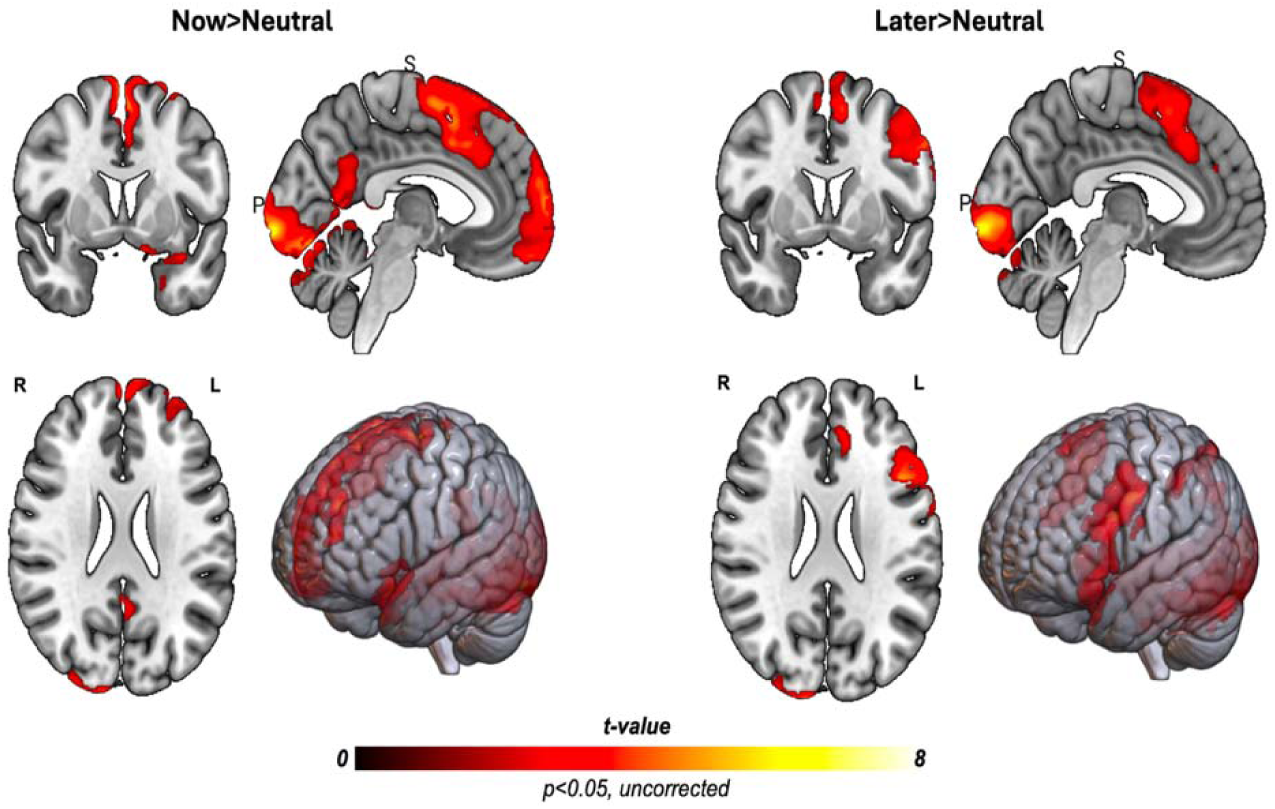
Brain response to ‘Now’ and ‘Later’ cues, relative to neutral cues: Left: fMRI BOLD signal in response to ‘Now’, relative to neutral cues, was elevated in the ventromedial and dorsomedial prefrontal, the posterior cingulate, and occipital cortices (voxel-wise threshold, p<0.05, Family-Wise Error cluster corrected). Right: fMRI BOLD signal in response to ‘Later’ relative to neutral cues was elevated in the mid cingulate, and occipital cortices (voxel-wise threshold, p<0.05, Family-Wise Error cluster corrected). There was also elevated BOLD signal within the left DLPFC, although this did not survive multiple comparison correction (voxel-wise threshold, p<0.05, cluster-level threshold, p<0.05 uncorrected).

**Supplemental Figure 2:**
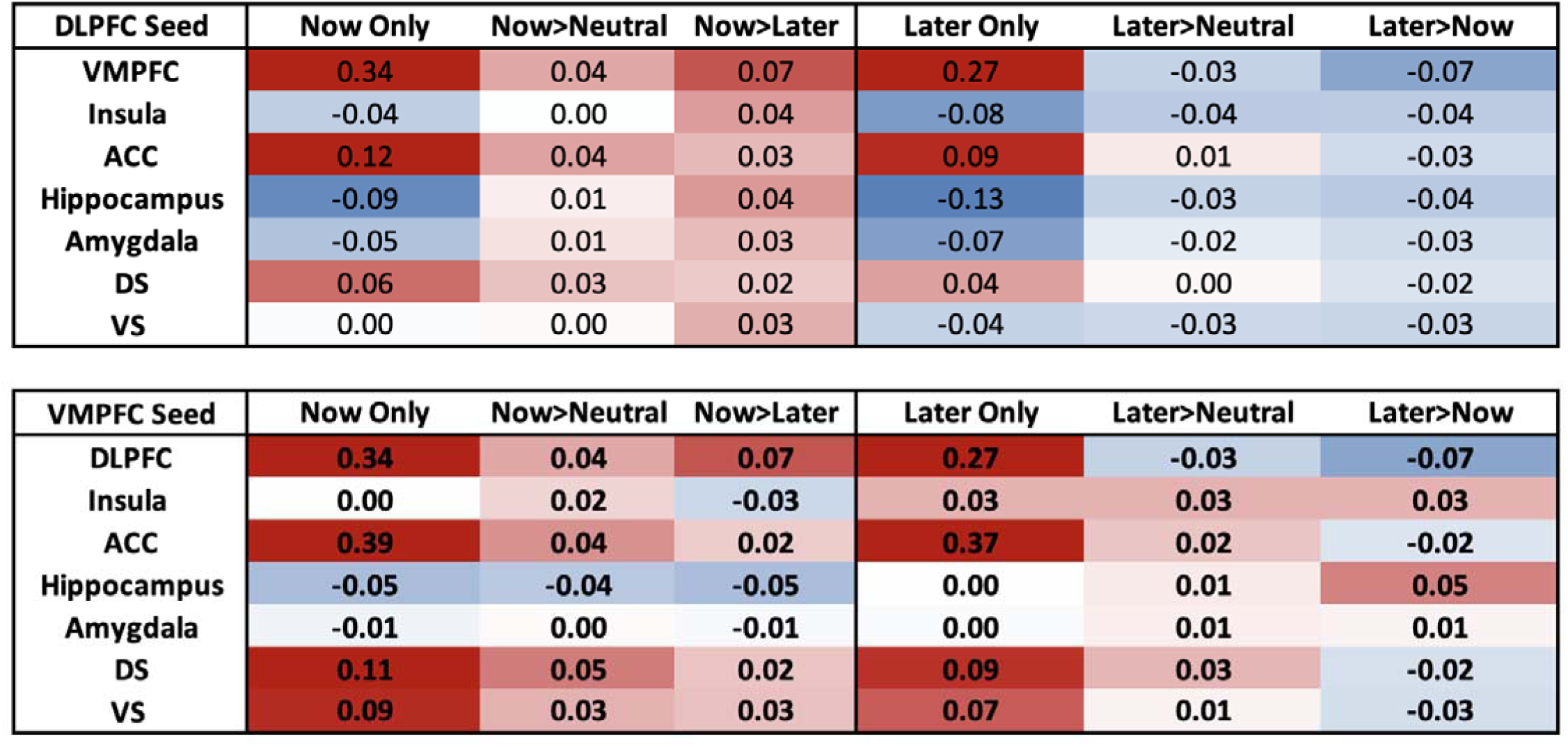
Functional connectivity patterns from the DLPFC and vMPFC during task conditions. Weighted ROI-to-ROI connectivity values from each seed region (DLPFC, top table; VMPFC, bottom table) during ‘Now’ and ‘Later’ cue conditions as well as contrasts with neutral and opposite conditions (e.g. Now > Later; Later > Now). Functional connectivity values during cues were generally highest fro the VMPFC to various downstream regions during Now cue conditions and contrasts, while connectivity values were generally lowest from the DLPFC to downstream regions during ‘Later’ cue conditions and contra ts.

**Supplemental-Table-1.** MNI coordinates of brain regions with elevated BOLD signal in response to ‘Now’, relative to ‘Later’ cues and in response to ‘Later’, relative to ‘Now’ cues. .

|  |  |  |  | MNI Coordinates |  |  |  |  |
| --- | --- | --- | --- | --- | --- | --- | --- | --- |
|  | L/R | Significant clusters | BA | X | Y | Z | Max t | Cluster-level p |
| 'NOW' relative to 'LATER' cues |  |  |  |  |  |  |  |  |
| Cluster 1,<br>k=3,996 | R | Postcentral gyrus | 4 | 22 | -28 | 76 | 6.78 | p<0.01 |
|  | R | Supplementary motor | 6 | 4 | -4 | 74 | 6.07 |  |
|  | L | Superior Parietal Lobule | 7 | -24 | -50 | 74 | 5.79 |  |
|  | L | Frontal superior gyrus | 6 | -20 | -4 | 76 | 5.76 |  |
|  | R | Postcentral gyrus | 4 | 16 | -30 | 80 | 5.57 |  |
|  | L | Postcentral gyrus | 4 | -16 | -30 | 76 | 5.46 |  |
|  | L | Postcentral gyrus | 1 | -24 | -30 | 76 | 5.31 |  |
|  | L | Middle frontal gyrus | 8 | -26 | 30 | 54 | 5.01 |  |
|  | R | Postcentral gyrus | 1 | 18 | -38 | 80 | 4.98 |  |
|  | L | Middle cingulate gyrus | 31 | -4 | -26 | 46 | 4.95 |  |
|  | L | Supplementary motor | 6 | -4 | -6 | 74 | 4.92 |  |
|  | L | Precentral gyrus | 6 | -2 | -14 | 74 | 4.89 |  |
|  | L | Posterior cingulate | 31 | -2 | -36 | 42 | 4.83 |  |
|  | L | Precentral gyrus | 6 | -12 | -16 | 78 | 4.82 |  |
|  | – | Precuneus | 7 | 0 | -48 | 66 | 4.79 |  |
|  | L | Postcentral gyrus | 5 | -24 | -36 | 74 | 4.79 |  |
| Cluster 2,<br>k=826 | R | dorsomedial PFC | 9 | 4 | 62 | 32 | 6.15 | p<0.01 |
|  | L | dorsomedial PFC | 10 | -4 | 66 | 22 | 4.96 |  |
|  | L | medial superior frontal gyrus | 10 | -2 | 60 | 30 | 4.94 |  |
|  | L | dorsomedial PFC | 10 | -2 | 62 | 26 | 4.88 |  |
|  | L | dorsomedial PFC | 9 | -4 | 60 | 34 | 4.59 |  |
|  | L | ventromedial PFC | 10 | -2 | 54 | -6 | 4.52 |  |
|  | L | frontal pole | 10 | -4 | 69 | 8 | 4.12 |  |
|  | L | anterior cingulate gyrus | 32 | -2 | 46 | 10 | 3.12 |  |
| Cluster 3,<br>k=2,101 | R | Thalamus | – | 8 | -12 | 18 | 5.91 | p<0.01 |
|  | L | Posterior cingulate | 30 | -6 | -50 | 6 | 5.58 |  |
|  | R | Lingual gyrus | 18 | 6 | -68 | -6 | 5.57 |  |
|  | L | Lingual gyrus | 18 | -10 | -44 | -4 | 5.42 |  |
|  | R | Lingual gyrus | 18 | 8 | -50 | 2 | 5.4 |  |
|  | R | Caudate | – | 8 | 10 | 12 | 5.01 |  |
|  | L | Lingual gyrus | 18 | -2 | -58 | 6 | 4.86 |  |
|  | – | Cerebellum | – | 0 | -68 | -6 | 4.82 |  |
|  | L | Cerebellum | – | -4 | -70 | -12 | 4.79 |  |
|  | R | Caudate | – | 10 | 18 | 12 | 4.79 |  |
|  | R | Caudate | – | 10 | 8 | 16 | 4.78 |  |
|  | L | Caudate | – | -10 | 2 | 16 | 4.75 |  |
|  | L | Cerebellum | – | -4 | -66 | -8 | 4.53 |  |
| 'LATER' relative to 'NOW' cues |  |  |  |  |  |  |  |  |
| Cluster 1,<br>k=2,572 | R | Angular gyrus | 39 | 48 | -44 | 24 | 7.93 | p<0.01 |
|  | R | Angular gyrus | 39 | 40 | -58 | 40 | 7.49 |  |
|  | R | Angular gyrus | 39 | 44 | -54 | 36 | 6.51 |  |
|  | R | Angular gyrus | 39 | 38 | -50 | 40 | 6.36 |  |
|  | R | Angular gyrus | 39 | 54 | -50 | 42 | 6.08 |  |
|  | R | Supramarginal gyrus | 40 | 48 | -48 | 42 | 5.94 |  |
|  | R | Angular gyrus | 39 | 48 | -46 | 38 | 5.48 |  |
|  | R | Superior temporal gyrus | 22 | 46 | -24 | -4 | 5.4 |  |
|  | R | Supramarginal gyrus | 40 | 38 | -40 | 38 | 5.05 |  |
|  | R | Angular gyrus | 39 | 40 | -58 | 28 | 4.92 |  |
|  | R | Angular gyrus | 39 | 40 | -62 | 16 | 4.84 |  |
|  | R | Middle occipital gyrus | 19 | 42 | -76 | 22 | 4.64 |  |
|  | R | Angular gyrus | 39 | 42 | -68 | 28 | 4.61 |  |
|  | R | Angular gyrus | 39 | 50 | -58 | 28 | 4.46 |  |
|  | R | Middle temporal gyrus | 39 | 54 | -48 | 12 | 3.97 |  |
| Cluster 2,<br>k=1,875 | R | Middle frontal gyrus | 8 | 38 | 4 | 40 | 6.54 | p<0.01 |
|  | R | DLPFC | 9 | 40 | 38 | 24 | 6.36 |  |
|  | R | Middle frontal gyrus | 6 | 36 | 6 | 52 | 6.04 |  |
|  | R | Broca's Area | 44 | 44 | 6 | 20 | 5.86 |  |
|  | R | DLPFC | 46 | 38 | 34 | 12 | 5.77 |  |
|  | R | Ventrolateral PFC | 10 | 40 | 50 | -12 | 5.62 |  |
|  | R | Broca's area | 44 | 40 | 8 | 22 | 5.61 |  |
|  | R | Superior frontal gyrus | 10 | 24 | 46 | 22 | 5.55 |  |
|  | R | Broca's Area | 44 | 34 | 8 | 26 | 5.48 |  |
|  | R | Ventrolateral PFC | 10 | 34 | 52 | 4 | 5.1 |  |
|  | R | Broca's Area | 44 | 38 | 6 | 26 | 4.96 |  |
|  | R | Broca's Area | 44 | 32 | 8 | 30 | 4.94 |  |
|  | R | Broca's Area | 44 | 34 | 8 | 30 | 4.89 |  |
|  | R | Lateral orbital gyrus | 47 | 36 | 46 | -12 | 4.76 |  |
|  | R | Precentral gyrus | 6 | 32 | -2 | 46 | 4.75 |  |
|  | R | Middle frontal gyrus | 10 | 46 | 50 | 8 | 4.75 |  |
| Cluster 3,<br>k=1,535 | L | Angular gyrus | 39 | -38 | -56 | 40 | 6.09 | p<0.01 |
|  | L | Middle temporal gyrus | 21 | -48 | -36 | -2 | 5.95 |  |
|  | L | Angular gyrus | 39 | -34 | -62 | 36 | 5.37 |  |
|  | L | Angular gyrus | 39 | -34 | -66 | 38 | 5.3 |  |
|  | L | Angular gyrus | 39 | -48 | -46 | 40 | 5.08 |  |
|  | L | Angular gyrus | 39 | -40 | -66 | 36 | 4.99 |  |
|  | L | Middle temporal gyrus | 21 | -64 | -46 | -2 | 4.98 |  |
|  | L | Middle temporal gyrus | 21 | -58 | -38 | -2 | 4.84 |  |
|  | L | Angular gyrus | 39 | -40 | -50 | 38 | 4.84 |  |
|  | L | Angular gyrus | 39 | -44 | -48 | 38 | 4.75 |  |
|  | L | Angular gyrus | 39 | -42 | -54 | 26 | 4.68 |  |
|  | L | Angular gyrus | 39 | -42 | -52 | 44 | 4.64 |  |
|  | L | Angular gyrus | 39 | -46 | -52 | 42 | 4.54 |  |
|  | L | Angular gyrus | 39 | -50 | -50 | 38 | 4.48 |  |
|  | L | Angular gyrus | 39 | -54 | -52 | 32 | 3.99 |  |

**Supplemental-Table-2.**
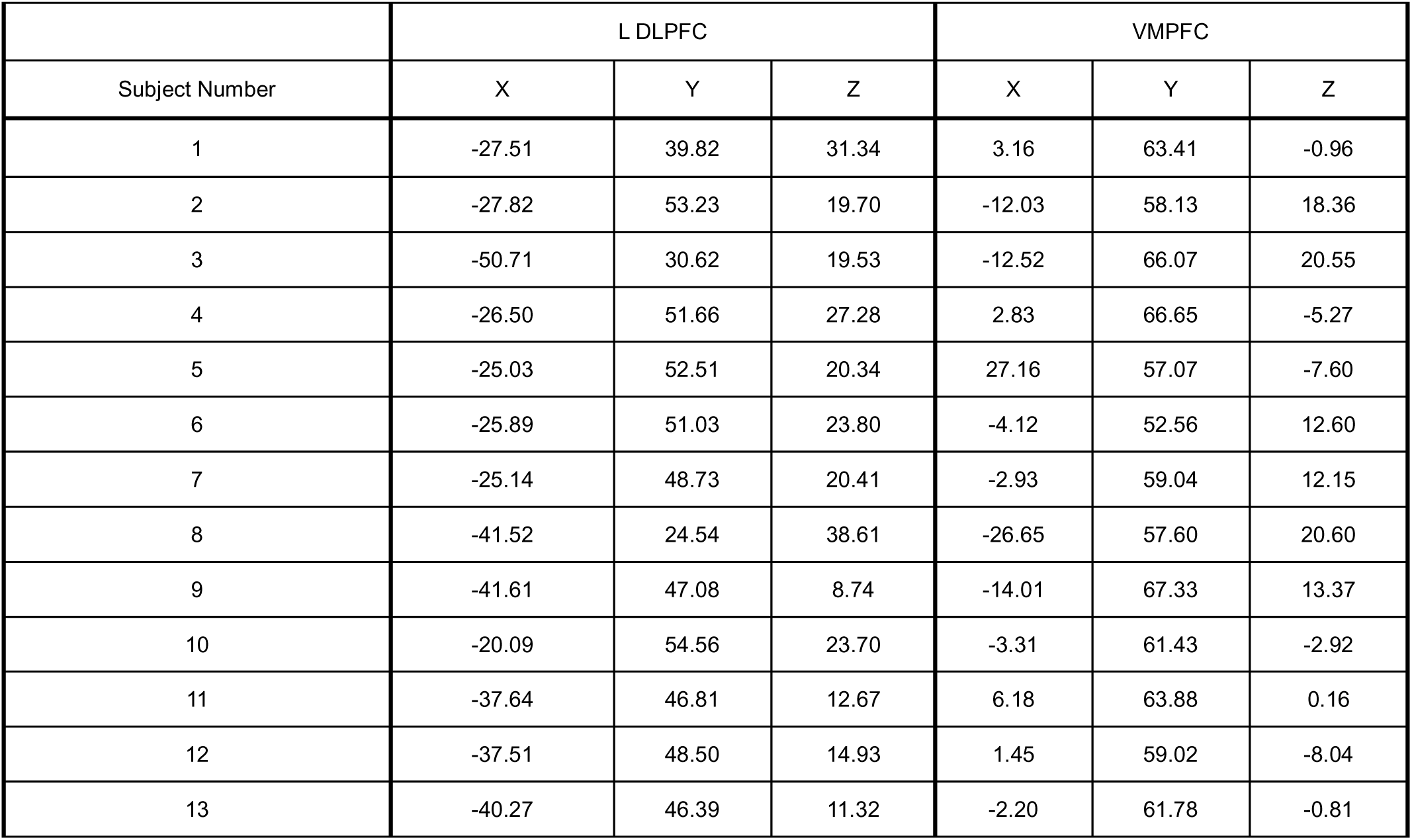
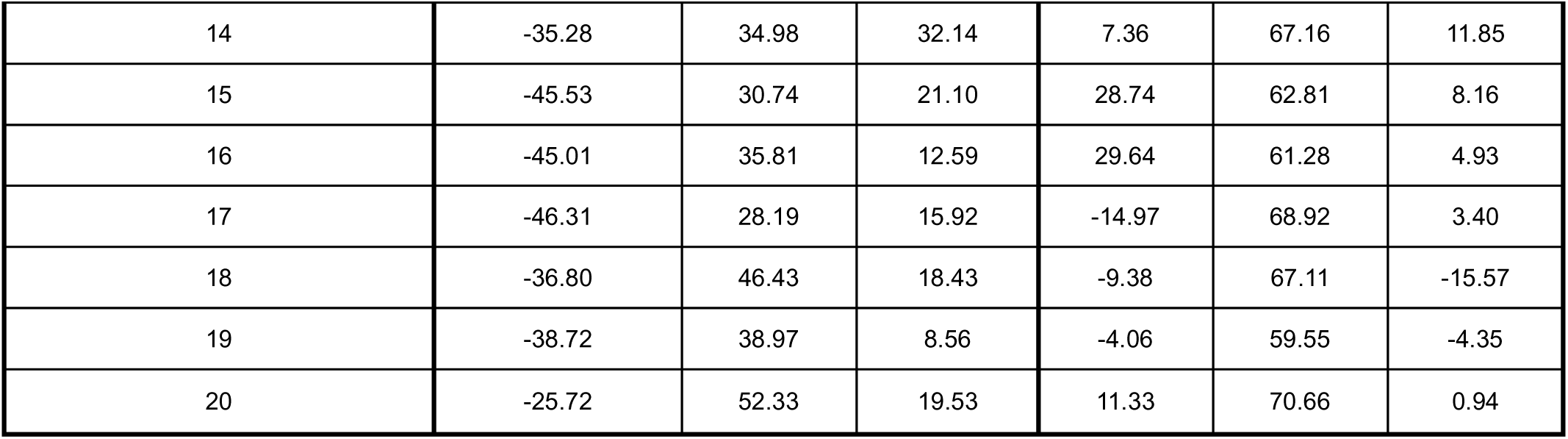
Target locations for both DLPFC and vmPFC in MNI space by participant:

|  | L DLPFC |  |  | VMPFC |  |  |
| --- | --- | --- | --- | --- | --- | --- |
| Subject Number | X | Y | Z | X | Y | Z |
| 1 | -27.51 | 39.82 | 31.34 | 3.16 | 63.41 | -0.96 |
| 2 | -27.82 | 53.23 | 19.70 | -12.03 | 58.13 | 18.36 |
| 3 | -50.71 | 30.62 | 19.53 | -12.52 | 66.07 | 20.55 |
| 4 | -26.50 | 51.66 | 27.28 | 2.83 | 66.65 | -5.27 |
| 5 | -25.03 | 52.51 | 20.34 | 27.16 | 57.07 | -7.60 |
| 6 | -25.89 | 51.03 | 23.80 | -4.12 | 52.56 | 12.60 |
| 7 | -25.14 | 48.73 | 20.41 | -2.93 | 59.04 | 12.15 |
| 8 | -41.52 | 24.54 | 38.61 | -26.65 | 57.60 | 20.60 |
| 9 | -41.61 | 47.08 | 8.74 | -14.01 | 67.33 | 13.37 |
| 10 | -20.09 | 54.56 | 23.70 | -3.31 | 61.43 | -2.92 |
| 11 | -37.64 | 46.81 | 12.67 | 6.18 | 63.88 | 0.16 |
| 12 | -37.51 | 48.50 | 14.93 | 1.45 | 59.02 | -8.04 |
| 13 | -40.27 | 46.39 | 11.32 | -2.20 | 61.78 | -0.81 |
| 14 | -35.28 | 34.98 | 32.14 | 7.36 | 67.16 | 11.85 |
| 15 | -45.53 | 30.74 | 21.10 | 28.74 | 62.81 | 8.16 |
| 16 | -45.01 | 35.81 | 12.59 | 29.64 | 61.28 | 4.93 |
| 17 | -46.31 | 28.19 | 15.92 | -14.97 | 68.92 | 3.40 |
| 18 | -36.80 | 46.43 | 18.43 | -9.38 | 67.11 | -15.57 |
| 19 | -38.72 | 38.97 | 8.56 | -4.06 | 59.55 | -4.35 |
| 20 | -25.72 | 52.33 | 19.53 | 11.33 | 70.66 | 0.94 |

## Methods

### Anatomical data preprocessing

For each participant, the T1-weighted (T1w) anatomical data were corrected for intensity non-uniformity (INU) with N4BiasFieldCorrection (Tustison et al. 2010), distributed with ANTs 2.3.3 (Avants et al. 2008, RRID:SCR_004757), and used as T1w-reference throughout the workflow. The T1w-reference was then skull-stripped with a *Nipype* implementation of the antsBrainExtraction.sh workflow (from ANTs), using OASIS30ANTs as target template. Brain tissue segmentation of cerebrospinal fluid (CSF), white-matter (WM) and gray-matter (GM) were then performed on the brain-extracted T1w using fast (FSL 5.0.9, RRID:SCR_002823, Zhang, Brady, and Smith 2001). Brain surfaces were reconstructed using recon-all (FreeSurfer 6.0.1, RRID:SCR_001847, Dale, Fischl, and Sereno 1999), and the brain mask estimated previously was refined with a custom variation of the method to reconcile ANTs-derived and FreeSurfer-derived segmentations of the cortical gray-matter of Mindboggle (RRID:SCR_002438, Klein et al. 2017). Volume-based spatial normalization to two standard spaces (MNI152NLin2009cAsym, MNI152NLin6Asym) was performed through nonlinear registration with antsRegistration (ANTs 2.3.3), using brain-extracted versions of both T1w reference and the T1w template. The following templates were selected for spatial normalization: *ICBM 152 Nonlinear Asymmetrical template version 2009c* [Fonov et al. (2009), RRID:SCR_008796; TemplateFlow ID: MNI152NLin2009cAsym], *FSL’s MNI ICBM 152 non-linear 6th Generation Asymmetric Average Brain Stereotaxic Registration Model* [Evans et al. (2012), RRID:SCR_002823; TemplateFlow ID: MNI152NLin6Asym],

### Functional data preprocessing

For each of the 3 BOLD runs per subject (across all tasks and sessions), the following preprocessing was performed. First, a reference volume and its skull-stripped version were generated using a custom methodology of *fMRIPrep*. A deformation field to correct for susceptibility distortions was estimated based on *fMRIPrep*’s *fieldmap-less* approach. The deformation field is that resulting from co-registering the BOLD reference to the same-subject T1w-reference with its intensity inverted (Wang et al. 2017; Huntenburg 2014). Registration was performed with antsRegistration (ANTs 2.3.3), and the process regularized by constraining deformation to be nonzero only along the phase-encoding direction and modulated with an average fieldmap template (Treiber et al. 2016). Based on the estimated susceptibility distortion, a corrected EPI (echo-planar imaging) reference was calculated for a more accurate co-registration with the anatomical reference. The BOLD reference was then co-registered to the T1w reference using bbregister (FreeSurfer) which implements boundary-based registration (Greve and Fischl 2009). Co-registration was configured with six degrees of freedom. Head-motion parameters with respect to the BOLD reference (transformation matrices, and six corresponding rotation and translation parameters) are estimated before any spatiotemporal filtering using mcflirt (FSL 5.0.9, Jenkinson et al. 2002). The BOLD time-series (including slice-timing correction when applied) were resampled onto their original, native space by applying a single, composite transform to correct for head-motion and susceptibility distortions. These resampled BOLD time-series will be referred to as *preprocessed BOLD in original space*, or just *preprocessed BOLD*. The BOLD time-series were resampled into standard space, generating a *preprocessed BOLD run in MNI152NLin2009cAsym space*. First, a reference volume and its skull-stripped version were generated using a custom methodology of *fMRIPrep*. Automatic removal of motion artifacts using independent component analysis (ICA-AROMA, Pruim et al. 2015) was performed on the *preprocessed BOLD on MNI space* time-series after removal of non-steady state volumes and spatial smoothing with an isotropic, Gaussian kernel of 6mm FWHM (full-width half-maximum). Corresponding “non-aggresively” denoised runs were produced after such smoothing. Additionally, the “aggressive” noise-regressors were collected and placed in the corresponding confounds file. Several confounding time-series were calculated based on the *preprocessed BOLD*: framewise displacement (FD), DVARS and three region-wise global signals. FD was computed using two formulations following Power (absolute sum of relative motions, Power et al. (2014)) and Jenkinson (relative root mean square displacement between affines, Jenkinson et al. (2002)). FD and DVARS are calculated for each functional run, both using their implementations in *Nipype* (following the definitions by Power et al. 2014). The three global signals are extracted within the CSF, the WM, and the whole-brain masks. Additionally, a set of physiological regressors were extracted to allow for component-based noise correction (*CompCor*, Behzadi et al. 2007). Principal components are estimated after high-pass filtering the *preprocessed BOLD* time-series (using a discrete cosine filter with 128s cut-off) for the two *CompCor* variants: temporal (tCompCor) and anatomical (aCompCor). tCompCor components are then calculated from the top 2% variable voxels within the brain mask. For aCompCor, three probabilistic masks (CSF, WM and combined CSF+WM) are generated in anatomical space. The implementation differs from that of Behzadi et al. in that instead of eroding the masks by 2 pixels on BOLD space, the aCompCor masks are subtracted a mask of pixels that likely contain a volume fraction of GM. This mask is obtained by dilating a GM mask extracted from the FreeSurfer’s *aseg* segmentation, and it ensures components are not extracted from voxels containing a minimal fraction of GM. Finally, these masks are resampled into BOLD space and binarized by thresholding at 0.99 (as in the original implementation). Components are also calculated separately within the WM and CSF masks. For each CompCor decomposition, the *k* components with the largest singular values are retained, such that the retained components’ time series are sufficient to explain 50 percent of variance across the nuisance mask (CSF, WM, combined, or temporal). The remaining components are dropped from consideration. The head-motion estimates calculated in the correction step were also placed within the corresponding confounds file. The confound time series derived from head motion estimates and global signals were expanded with the inclusion of temporal derivatives and quadratic terms for each (Satterthwaite et al. 2013). Frames that exceeded a threshold of 0.5 mm FD or 1.5 standardised DVARS were annotated as motion outliers. All resamplings can be performed with *a single interpolation step* by composing all the pertinent transformations (i.e. head-motion transform matrices, susceptibility distortion correction when available, and co-registrations to anatomical and output spaces). Gridded (volumetric) resamplings were performed using antsApplyTransforms (ANTs), configured with Lanczos interpolation to minimize the smoothing effects of other kernels (Lanczos 1964). Non-gridded (surface) resamplings were performed using mri_vol2surf (FreeSurfer).

Many internal operations of *fMRIPrep* use *Nilearn* 0.6.2 (Abraham et al. 2014, RRID:SCR_001362), mostly within the functional processing workflow. For more details of the pipeline, see <u>the section corresponding to workflows in *fMRIPrep*</u>’s documentation.

### Task-Based Functional Connectivity

Task-Based functional connectivity was calculated using the Conn toolbox (version 20.b). vmPFC and LDLPFC were set as seed regions. A weighted general linear model and region of interest (ROI)-to-ROI analysis were performed. To evaluate change within specific ROIs, we extracted connectivity values (Fischer’s z-scores) to an a priori network of regions^5,24^, which include: the stimulation targets, vmPFC and DLPFC, the bilateral anterior cingulate cortex, left and right dorsal striatum, left and right ventral striatum, the left and right insula, and left and right hippocampus. Because we collected a behavioral measure of withdrawal at a time-point of ad libidum use and at 24-hours of abstinence, we also performed an exploratory correlational analysis between left and right amygdala and withdrawal sensitivity, necessitating extraction of the average BOLD signal in the left and right amygdala.

## Data Availability

All data produced in the present study are available upon reasonable request to the authors

## Acknowledgements

The authors would like to thank the many contributors to this work including Hua Wu, Lauren Crowe, Saron Hunegnaw, Noriah Johnson, and Ethan Makarewycz.

## Declaration of Interests

AG is a current employee of Magnus Medical, Inc. ALM has served as a consultant for Indivior and received grant funding from Pleo Pharma. GLS has collaborated with MagVenture and MECTA as part of investigator-initiated trials, consults for and has equity in the company Trial Catalyst LLC, and has provided consultation to Indivior. All other authors report no biomedical financial interests or potential conflicts of interest.

## Primary Funding

This work was supported by the Stanford Neurochoice initiative, The Taube Foundation, the National Institute of Alcohol Abuse and Alcoholism (McCalley, K99AA031508), and the National Institute of Drug Abuse (Froeliger, UG3DA048510, Mcrae-Clark, K24DA038240, and Sahlem, K23DA043628). Work on “The Development and Validation of Neural Targets in Opioid Use Disorder for Use Across Addictions” was supported by Wellcome Leap as part of the Untangling Addiction program.

## Author Contributions

AG, WS, AA, BW, ALM, NRW, BF, and GLS designed the experiment. AG, WS, AA, BW, SN, SA and GLS conducted the experiment. AG, DM, WS, AA, BW, BK, JPK, and GLS performed the analysis. AG, DM, WS, AA, BK, JPK, and GLS wrote the first draft of the manuscript. All authors critically reviewed and edited the manuscript.

## Clinicaltrials.gov identifiers

NCT05720312

## Notes

### Clinical Trial

NCT05720312

### Author Declarations

The parent trial was registered with clinicaltrials.gov, approved by the Stanford University Institutional Review Board, and was conducted in accordance with the principles outlined in the Declaration of Helsinki. All participants signed informed consent prior to beginning study procedures.

